# Human serum from SARS-CoV-2 vaccinated and COVID-19 patients shows reduced binding to the RBD of SARS-CoV-2 Omicron variant

**DOI:** 10.1101/2021.12.10.21267523

**Authors:** Maren Schubert, Federico Bertoglio, Stephan Steinke, Philip Alexander Heine, Mario Alberto Ynga-Durand, Fanglei Zuo, Likun Du, Janin Korn, Marko Milošević, Esther Veronika Wenzel, Henrike Maass, Fran Krstanović, Saskia Polten, Marina Pribanić-Matešić, Ilija Brizić, Antonio Piralla, Fausto Baldanti, Lennart Hammarström, Stefan Dübel, Alan Šustić, Harold Marcotte, Monika Strengert, Alen Protić, Qiang Pan-Hammarström, Luka Čičin-Šain, Michael Hust

**Affiliations:** Technische Universität Braunschweig, Institut für Biochemie, Biotechnologie und Bioinformatik, Abteilung Biotechnologie, Spielmannstr. 7, 38106 Braunschweig, Germany; Helmholtz Centre for Infection Research, Department of Viral Immunology, Inhoffenstr. 7, 38124 Braunschweig, Germany; Department of Biosciences and Nutrition, Karolinska Institutet, Huddinge, Sweden; Abcalis GmbH, Science Campus Braunschweig-Süd, Inhoffenstr. 7, 38124 Braunschweig, Germany; Department of Anesthesiology, Reanimation, Intensive Care and Emergency Medicine, Faculty of Medicine, University of Rijeka, Rijeka, Croatia; Center for Proteomics, Faculty of Medicine, University of Rijeka, Rijeka, Croatia; Molecular Virology Unit, Microbiology and Virology Department, Fondazione IRCCS Policlinico San Matteo, Pavia, Italy; Department of Clinical, Surgical, Diagnostic and Paediatric Sciences, University of Pavia, Pavia, Italy; Department of Epidemiology, Helmholtz Centre for Infection Research, Inhoffenstr. 7, 38124 Braunschweig, Germany; Centre for Individualised Infection Medicine (CIIM), a joint venture of Helmholtz Centre for Infection Research and Medical School Hannover

## Abstract

The COVID-19 pandemic is caused by the betacoronavirus SARS-CoV-2. In November 2021, the Omicron variant was discovered and classified as a variant of concern (VOC). Omicron shows substantially more mutations in the spike protein than any previous variant, mostly in the receptor binding domain (RBD). We analyzed the binding of the Omicron RBD to the human ACE2 receptor (hACE2) and the ability of human sera from COVID-19 patients or vaccinees in comparison to Wuhan, Beta or Delta RBDs variants.

All RBDs were produced in insect cells. RBD binding to hACE2 was analyzed by ELISA and microscale thermophoresis (MST). Similarly, sera from 27 COVID-19 patients, 58 fully vaccinated individuals and 16 booster recipients were titrated by ELISA on the fixed RBDs from the original Wuhan strain, Beta, Delta and Omicron VOC.

Surprisingly, the Omicron RBD showed a weaker binding to ACE2 compared to Beta and Delta, arguing that improved ACE2 binding is not a likely driver of Omicron evolution. Serum antibody titers were significantly lower against Omicron RBD compared to the original Wuhan strain. However, a difference of 2.5 times was observed in RBD binding while in other studies the neutralization of Omicron SARS-CoV-2 was reduced by a magnitude of 10x and more. These results indicate an immune escape focused on neutralizing antibodies.

The reduced binding of sera to Omicron RBD adds evidence that current vaccination protocols may be less efficient against the Omicron variant.

## Introduction

SARS-CoV-2 is the etiological agent of the severe pneumonia COVID-19 (coronavirus disease 2019) *(1, 2)*. A new variant B.1.1.529 of the betacoranavirus SARS-CoV-2 was identified in late November 2021 and has rapidly been classified as a variant of concern (VOC) by the WHO and named Omicron *(3)*. The Omicron variant shows a high number of mutations in the SARS-CoV-2 spike protein in comparison to the previously described VOCs Alpha *(4)*, Beta (B.1.351) *(5)*, Gamma (P.1) *(6)* and the currently dominating Delta variant (B.1.617.2) *(7)*. The first sequenced Omicron variant (GISAID accession ID EPI_ISL_6913995, collection date 2021-11-08, South Africa) contains a total of 36 mutations compared to the original Wuhan strain and include 29 amino acid (aa) changes, six aa deletions and one aa insertion. Fifteen of these mutations are concentrated in the N-terminal receptor binding domain (RBD) of the spike protein which binds to the human zinc peptidase angiotensin-converting enzyme 2 (ACE2) for cell entry *(8, 9)*.

Importantly, the RBD is targeted by more than 90% of the neutralizing serum antibodies, making it the most relevant target for SARS-CoV-2 neutralization *(10, 11)*. Consequently, the majority of therapeutic antibodies for the treatment of COVID-19 are designed to interact with this part of the SARS-CoV-2 spike protein *(12, 13)*. This indicates that the Omicron variant may bind with a different affinity to the cell receptor, altering its propagation characteristics. Alternatively, the mutations may help the virus to escape the immune recognition by antibodies, facilitating viral spread in a seropositive population. Finally, both phenomena may occur in parallel.

While initial studies have shown a severe reduction in serum neutralizing capacity of vaccinated and convalescent patients against the Omicron variant *(14)*, it is unclear to which extent the RBD domain mutations contribute to this loss in neutralization activity. Additionally, while several mutations present in Omicron are predicted to increase ACE2 binding affinity, others are predicted to reduce its affinity *(15)*.

In this work, the binding of ACE2 to the new Omicron RBD was determined in comparison to the original Wuhan strain and the Beta and Delta variants in a biological experimental model. Moreover, we tested the binding of human sera from COVID-19 hospitalized patients or fully vaccinated persons with BNT162b2 or Ad26.COV2.S, as well as boost vaccinated persons, to the RBD of the original Wuhan strain, the Beta, the Delta and the Omicron VOC.

## Results

### The Omicron RBD shows a slightly reduced binding to ACE2

The RBD of the original Wuhan strain, the Beta, Delta and the Omicron variants were produced in insect cells and purified by IMAC and SEC. The quality of the recombinant proteins was analyzed by SDS-PAGE (Supplementary Figure 1). The production yield of Omicron RBD was tendentially higher than the yield of Wuhan RBD. All RBDs were immobilized on plates and binding of the soluble receptor ACE2 was analyzed by ELISA (Figure 1). The Omicron RBD showed a slightly reduced binding to ACE2 (EC_50_ 150 ng/mL) compared to the Wuhan strain RBD (EC_50_ 120 ng/mL) and a reduced binding to Beta (EC_50_ 89 ng/mL) and Delta RBD (EC_50_ 89 ng/mL). The affinities were also measured by microscale thermophoresis (MST) (Table 1, Supplementary Figure 2). The measured affinity for the Omicron RBD was notably lower compared to Beta and Delta.

**Table 1:** RBD - ACE2 affinity measured by MST. All experiments were performed in titration in triplicates and analyzed by the MO Affinity Analysis software (NanoTemper) by Hill fit.

| ***RBD*** | ***EC50 (nM)*** | ***EC50 confidence (nM)*** |
| --- | --- | --- |
| Wuhan strain | 40.7 | 1.8 |
| Beta | 35.2 | 1.7 |
| Delta | 33.6 | 2.2 |
| Omicron | 42.3 | 1.6 |

**Figure 1:**
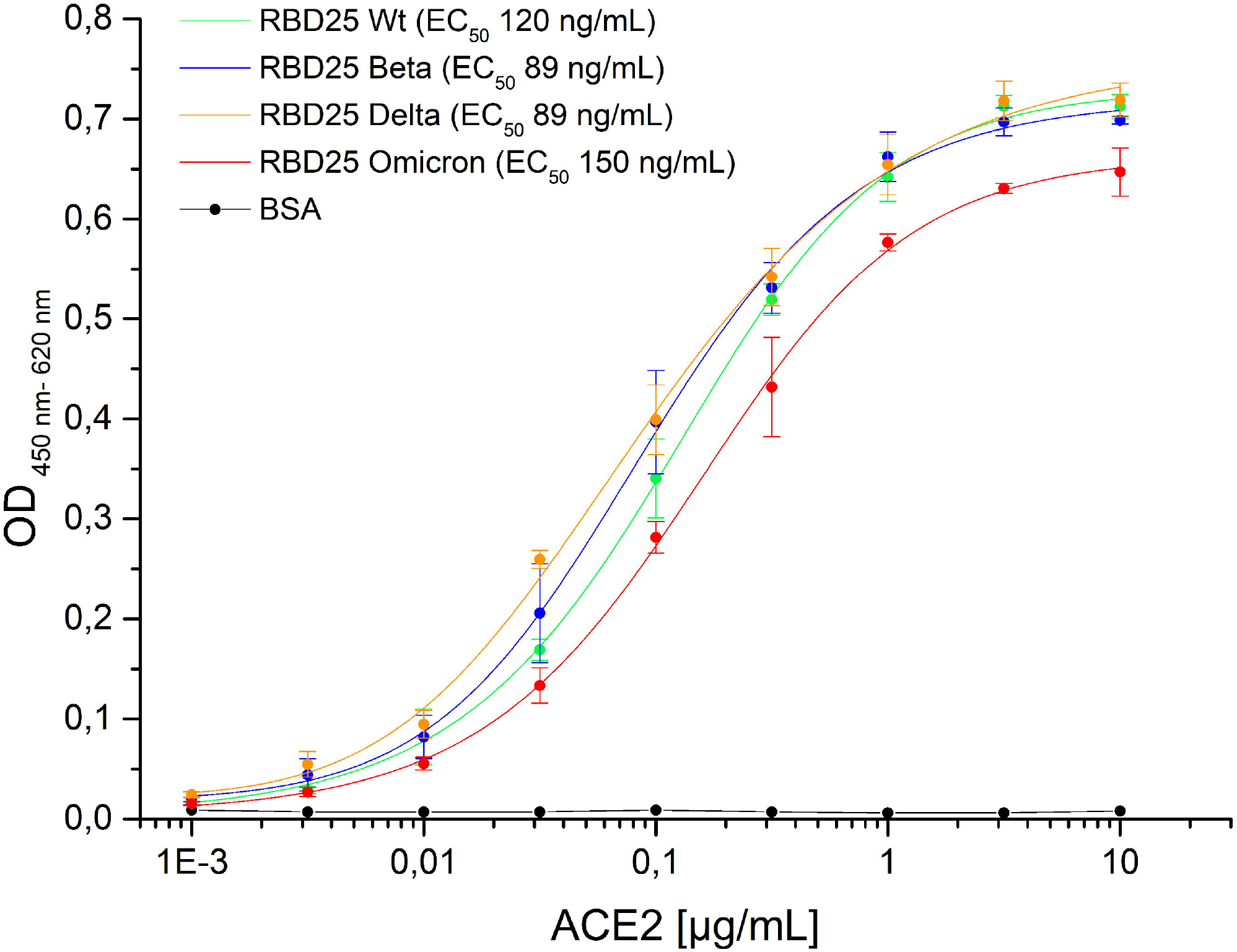
RBD variants binding to human ACE2. 300 ng/well immobilized Wuhan wt, Beta, Delta or Omicron RBD were detected with human ACE (fusion protein with human Fc part) in titration ELISA. BSA was used as negative control. Experiments were performed in triplicates and mean values are given. EC_50_ were calculated with OriginPro Version 9.1, fitting to a five-parameter logistic curve.

### Human sera of COVID-19 patients and vaccinated persons show a reduced binding to Omicron RBD

The binding of human sera from hospitalized COVID-19 patients (Figure 2A), from people fully vaccinated with BNT162b2 (Corminaty) (7-43 days after second immunization) (Figure 2B), Ad26.COV2.S (Janssen COVID-19 Vaccine) (14-33 days after immunization) (Figure 2C), mRNA1273 (Spikevax) (5-55 days after second immunization) (Figure 2D), or from mRNA vaccine boost-vaccinated persons (5-49 days after boost vaccination, first immunization 2xBNT162b2 or 1xAd26.COV2.S) (Figure 2E) was analyzed by ELISA on Wuhan, Delta, Beta and Omicron RBD. A direct comparison of the binding to Omicron RBD of all four serum groups is given in Figure 2F.

**Figure 2:**
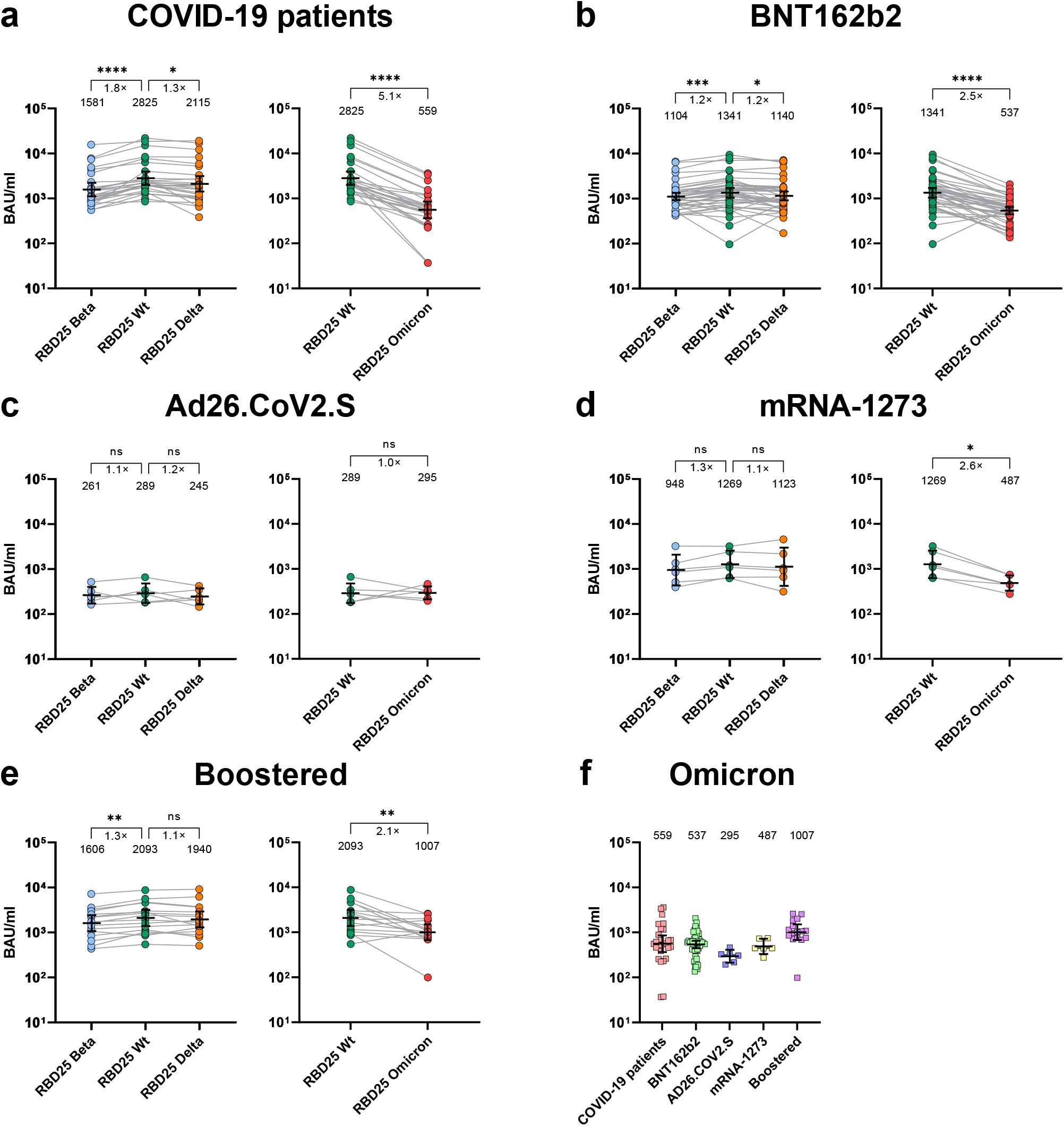
Human serum binding to SARS-CoV-2 Wuhan original strain, Beta, Delta and Omicron RBD. (A) ELISA using sera from hospitalized COVID-19 patients. (B) ELISA using sera from 2xBNT162b2 vaccinated persons (7-52 days after 2nd immunization). (C) ELISA using sera from 1xAd26.COV2.S vaccinated (14-33 days 1st immunization). (D) ELISA using sera from 2xBNT162b2 or Ad26.COV2.S vaccinated and boosted with BNT162b2 or mRNA-1273 (5-49 days after 3rd or in case of Ad26.COV2.S 2nd immunization). (E) ELISA using sera from 2xBNT162b2 or Ad26.COV2.S vaccinated and boosted with BNT162b2 or mRNA-1273 (5-49 days after 3rd or in case of Ad26.COV2.S 2nd immunization) binding to the Omicron variant. This is a rearranged representation of the data presented in (A-E). The ELISAs were performed as single point titrations. EC_50_ were calculated by DataGraph Version 4.7.1, fitting to a four-parameter logistic curve and expressed as relative potency in respect to an internal calibrant, for which the Binding Antibody Unit (BAU) was calculated using the WHO International Standard 20/136 titrated on Wuhan wt as reference. The geometrical mean values and the 95% CI are given in the graphs. The graphics and statistical analysis were made with Graphpad Prism 9.1.

The sera of COVID-19 patients, BNT162b2 and mRNA-1273 showed a highly significant reduction in binding to Omicron RBD in a non-parametric pairwise analysis. This reduction was more pronounced than the one observed against Beta or Delta RBD binding assays. In the booster group the binding to original Wuhan strain, Beta and Delta RBDs was nearly equal, but significantly reduced to Omicron RBD. Ad26.COV2.S group showed very low binding to all RBDs tested at all, suggesting a clearly weak immunogenicity of this vaccine formulation.

Boost recipients had a lower serum antibody binding to Omicron RBD compared to Wuhan, Beta and Delta (Figure 2E). On the other hand, the boost increased the amount of anti-Omicron RBD antibodies in comparison to the sera of fully vaccinated people (Figure 2F).

## Discussion

RBD-ACE2 interaction is a prerequisite for SARS-CoV-2 viral entry *(8, 9, 16)*. The binding of the Omicron RBD to the ACE2 receptor appears to be reduced in our settings compared to the currently dominant Delta variant, both in an ELISA assay as well as by affinity measurement using MST. The measured affinities (40.7 nM for Wuhan RBD) were in the same range as the ACE2 affinities determined previously by surface plasmon resonance (44.2 nM) for the same RBD variant *(17)*. Several Omicron RBD mutations are assumed to increase the binding to ACE2: G339D, S477N, T478K, Q493K, N501Y, others are neutral: S371L, S373P, G446S, E484A, Q493R, Q498R or are assumed to reduce the binding to ACE2: S375F, K417N, G496S, Y505H according yeast display experiments performed by Starr et al. *(16)*. Hence, some bioinformatic modelers predicted an increase in the ACE2 binding affinity of Omicron RBD *(18)* while other models rejected this scenario *(19, 20)* and stated: “the Q493R/K mutations, in a combination with K417N and T478K, dramatically reduced the S1 RBD binding by over 100 folds” *(19)*. However, the latter considerations were based on *in silico* modeling. Thus, RBD-ACE2 interaction involving a heavily mutated RBD, such as the one of Omicron VOC, may deviate from predictions and requires empirical biochemical testing. To our knowledge, this is the first comprehensive empirical analysis of Omicron RBD binding efficacy to the ACE2 receptor. Surprisingly, the binding of Omicron RBD to human ACE2 was not increased, but rather decreased, especially when compared to Beta and Delta. A recent study with Omicron pseudotyped viruses has indicated that ACE2 remains necessary for cell entry of this VoC *(21)*. However, the decrease in RBD binding does not necessarily translate into reduced infectivity, as infectivity and replication is also defined by proteolytic spike processing, fusion efficacy and RNA replication efficiency, just to name a few mechanisms. Nevertheless, our results argue that increased binding to the hACE2 receptors may be an unlikely cause of rapid Omicron spread. One has also to consider that we utilized in our work the originally available sequence with a Q493K mutation, whereas Q493R sequences have been published since. According to Starr et al. *(16)*, the K mutation has a higher affinity as the R mutation in *in vitro* binding studies. On the other hand, others have calculated an increased binding to ACE2 for the Q493R mutation in docking studies *(15)*.

Importantly, RBD mutations may also lead to immune escape *(22)*. The humoral immune answer is a key factor for the anti-viral defense *(23)* and the RBD is the main target of neutralizing antibodies *(10, 11, 24)*. The very low RBD binding in the Ad26.COV2.S compared to the COVID-19 patient group or the BNT162b2 vaccinated group is in accordance with the former results *(25)* but impaired definite conclusions on Omicron immune escape upon Ad26.COV2.S vaccination. The reduced binding of sera from COVID-19 patient and BNT162b2 groups to the Omicron RBD was in accordance with current results describing a highly reduced neutralization of the Omicron variant by human sera from vaccinated persons *(14, 21, 26)*. This also shows, that the Omicron mutations mainly reduce the SARS-CoV-2 neutralization but not the RBD binding in the same measure indicating an immune escape focused on neutralization. This reduced infectivity seems to be compensated by increased viral replication in ex vivo explant cultures of human bronchus *(27)*.

Sera from boosted vaccine recipients showed a significant reduction in serum titers against the Omicron variant, but not against the Beta or Delta variant. This observation, together with data on reduced Omicron-RBD binding to ACE2, argues that Omicron-RBD mutations may have arisen primarily to escape immune recognition, rather than to facilitate viral spread. This strengthens the case for the rapid development of novel Omicron directed vaccine formulations. While we have measured significant decrease of serum binding to Omicron RBD, the boost recipients had still a higher anti-Omicron RBD titer than fully vaccinated patients. This highlights the importance of boost immunization as observed in other studies *(14, 21)*.

The reduced binding of the sera from patient, vaccinated and boost-vaccinated individuals to the Omicron RBD is a snapshot of the current situation. According to the sequencing data deposited as GISAID (https://www.gisaid.org/) and the analysis on Outbreak.info *(28)*, the frequency of the 15 aa mutations in the RBD is very dynamic, e.g. K417N described for the initial Omicron variant is occurs in ~35% of all sequenced Omicron variants, S477N, T478K and E484A is ~47%, N501Y is in ~85% of the sequences (status 2021-12-14, 2146 sequences). The K417N mutation is a key mutation also in the Beta variant, the T478K mutation in the Delta variant and N501Y in the Alpha, Beta and Gamma variants *(29)*. All of these mutations may contribute to both ACE2 binding efficacy and immune escape. Therefore, Omicron variants with alternative mutations might evolve in the near future and alter the antibody recognition and/or the ACE2 binding efficacy. More comprehensive studies of various subvariants in the Omicron family may shed the light on their biochemical and immunological properties and understand the potential for future SARS-CoV-2 evolution.

## Material & Methods

### Serum samples

Blood samples were obtained from hospitalized patients with severe symptoms from the second (pre-alpha) and third (alpha variant) pandemic wave in Croatia or from vaccinated people as indicated. While all voluntary donors were informed about the project and gave their consent for the study, consent requirement was waived by the ethical committee in Rijeka for patients in intensive care where sampling was a part of routine diagnostics. The sampling was performed in accordance with the Declaration of Helsinki. The donors included adults of both sex. The first WHO International Standard for anti-SARS-CoV-2 immunoglobulin (NIBSC code: 20/136) was used as positive control serum and pre-pandemic negative control sera were provided by the LADR Braunschweig. Approval was given from the ethical committee of the Technische Universität Braunschweig (Ethik-Kommission der Fakultät 2 der TU Braunschweig, approval number FV-2020-02). The study in Croatia was approved by the Ethics committee of the Rijeka Clinical Hospital Center (2170-29-02/1-20-2). The study in Italy was performed under the approval of the Institutional Review Board of Policlinico San Matteo (protocol number P_20200029440). The study in Sweden was approved by the ethics committee in Stockholm (Dnr 2020-02646).

Details about study participants are shown in Table 2.

**Table 2:**
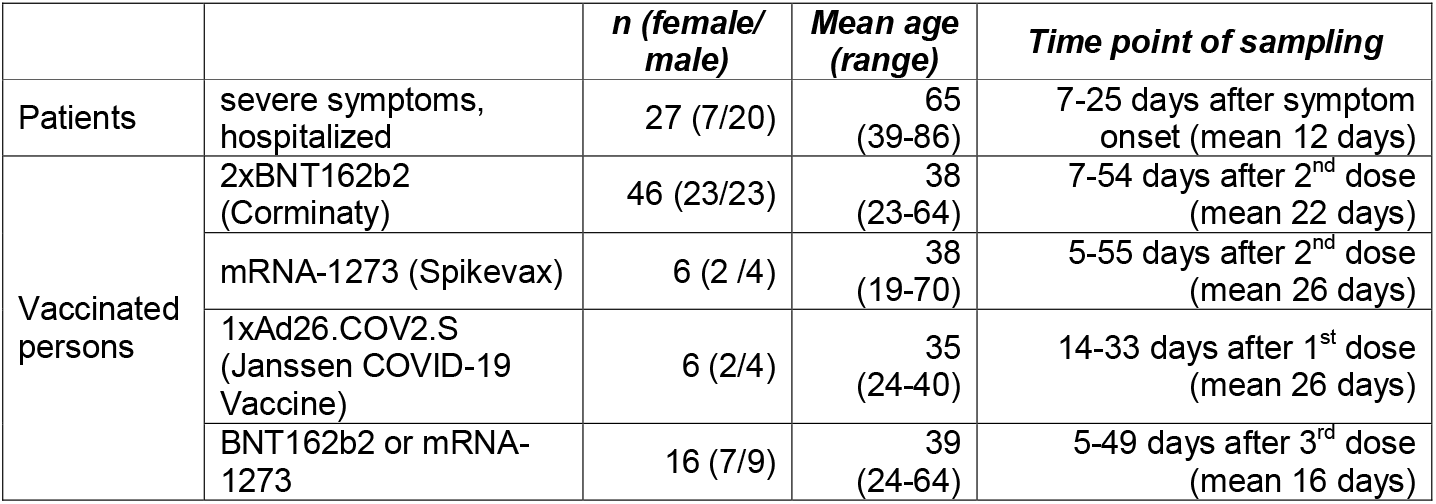
Used human serum samples in this study.

#### Construction of the expression vectors

All sequences of the RBD variants (319-541 aa of GenBank: MN908947) were inserted in a *Nco*I/*Not*I compatible variant of the OpiE2 expression vector *(30)* containing a N-terminal signal peptide of the mouse Ig heavy chain and a C-terminal 6xHis-tag. Single point mutations to generate the Beta and Delta variants of RBD were inserted into the original Wuhan strain through site-directed mutagenesis using overlapping primers according to Zheng et al. with slight modifications: S7 fusion polymerase (Mobidiag) with the provided GC buffer and 3% dimethyl sulfoxide was used for the amplification reaction. The RBD omicron variant was ordered as GeneString from GeneArt (Thermo Fisher) according to EPI_ISL_6590608 (partial RBD Sanger sequencing from Hong Kong), EPI_ISL_6640916, EPI_ISL_6640919 and EPI_ISL_6640917 including Q493K which was corrected later to Q493R. Table 3 gives an overview about the used variants.

**Table 3:** RBD variants used in this study (319-541 of GenBank: MN908947)

|  |  |  |
| --- | --- | --- |
| RBD wt | Original Wuhan | - |
| RBD beta | B.1.351 | K417N, E484K, N501Y |
| RBD delta | B.1.617.2 | L452R, T478K |
| RBD omicron | B.1.1.529 | G339D, S371L, S373P, S375F, K417N, N440K, G446S, S477N, T478K, E484A, Q493K, G496S, Q498R, N501Y, Y505H |

### Expression and purification of the RBD variants

The different RBD variants were produced in the baculovirus-free High Five cell system *(31)* and purified as described before *(32)*. Briefly, High Five cells (Thermo Fisher Scientific) were cultivated at 27°C, 110-115 rpm in EX-CELL 405 media (Sigma Aldrich) at a cell density between 0.3– 5.5×10^6^ cells/mL. On the day of transfection, cells were centrifuged and resuspended in fresh media to a density of 4×10^6^ cells/mL before 4 µg expression plasmid/mL and 16 µg/mL of linear PEI 40 kDa (Polysciences) was pipetted directly into the cell suspension. After 4-24 h, cells were supplemented with fresh media to dilute the cells ~1×10^6^ cells/mL and 48 h after transfection, culture volume was doubled. Cell supernatant was harvested four to five days after transfection by a two-step centrifugation (4 min at 180 xg and 20 min at >3,500 xg) and then 0.2 µm filtered for purification. Immobilized Metal Ion Affinity Chromatography (IMAC) His tag purification of insect cell supernatant was performed with a HisTrap excel column (Cytiva) on Äkta system (Cytiva) according to the manufacturer’s manual. In a second step, the RBD domains were further purified by size exclusion chromatography (SEC) by 16/600 Superdex 200 kDa pg column (Cytiva).

### Expression and purification of ACE2-hFc

The extracellular domain of ACE2 receptor (GenBank NM_021804.3) was produced in pCSE2.6-hFc expression vector in Expi293F cells (Thermo Fisher Scientific) as described before (Bertoglio et al., 2021b). In brief, Expi293F cells were cultivated at 37°C, 110 rpm, and 5% CO_2_ in Gibco FreeStyle F17 expression media (Thermo Fisher Scientific) supplemented with 8 mM Glutamine and 0.1% Pluronic F68 (PAN Biotech). For transfection 1 μg DNA and 5 μg of 40 kDa PEI (Polysciences) per mL transfection volume were diluted separately in 5 transfection volume and than mixed for formation of complexes (20-30 min). Afterwards, PEI:DNA complexes were added to 1.5-2×10^6^ cells/mL. Forty-eight hours later, the culture volume was doubled by feeding HyClone SFM4Transfx-293 media (GE Healthcare) supplemented with 8 mM Glutamine and HyClone Boost 6 supplement (GE Healthcare) with 10% of the end volume. One week after transfection, supernatant was harvested by 15 min centrifugation at 1,500 ×g. Purification was performed on 1 mL HiTrap Fibro PrismA (Cytiva) column on Äkta go (Cytiva) according to the manufacturer’s manual.

### ACE2 titration ELISA on RBD

ACE2 binding to the produced RBD variant antigens was analyzed in ELISA in triplicates where 300 ng RBD per well was immobilized on a Costar High binding 96 well plate (Corning, Costar) at RT for 1 h. Next, the wells were blocked by 330 μL 2% MPBST (2% (w/v) milk powder in PBS; 0.05% Tween20) for 1 h at RT and then washed 3 times with H_2_O and 0.05% Tween20 (BioTek Instruments, EL405). ACE2-hFc was titrated from 0.01 mg/mL down to 1 ng/mL and incubated 1 h at RT prior to another 3x times washing step. Detection was performed by goat-anti-hIgG(Fc) conjugated with HRP (1:70,000, A0170, Sigma) and visualized with tetramethylbenzidine (TMB) substrate (20 parts TMB solution A (30 mM potassium citrate; 1% (w/v) citric acid (pH 4.1)) and 1 part TMB solution B (10 mM TMB; 10% (v/v) acetone; 90% (v/v) ethanol; 80 mM H_2_O_2_ (30%)) were mixed). After addition of 1 N H_2_SO_4_ to stop the reaction, absorbance at 450 nm with a 620 nm reference wavelength was measured in an ELISA plate reader (BioTek Instruments, Epoch). EC_50_ were calculated with OriginPro Version 9.1, fitting to a five-parameter logistic curve.

### Affinity measurement by microscale thermophoresis

The affinity measurements were performed as described before *(33)*. In brief, ACE2-hFc was labeled by the Protein Labeling Kit RED-NHS 2nd Generation (NanoTemper) according to the manufacturer’s protocol. A DOI of <3 was achieved and 10 nM of the labeled ACE2-hFc was applied in the measurements. Titration of the RBD variants was done by Precision XS microplate sample processor (BioTek) in 384 well plates. Measurement was performed in Monolith (Nanotemper) using Monolith NT.Automated Capillary Chips (NanoTemper). The Excitation-Power was set to 40% and MST-Power to medium. The timeframe of 0.5 s up to 1.5 s was chosen to analyze the data by the MO Affinity Analysis software (NanoTemper) by Hill fit. For all RBD variants a signal response above 18 and a signal to noise above 40 was obtained.

### Serum titration ELISA

For titration ELISA, sera were diluted 1:100 to 1: 9.19×10^7^ in 384 well microtiter plates (Greiner Bio-One) coated with 30 ng/well of the respective RBD variant. In addition, all sera were also tested at the lowest dilution (1:100) for determination of unspecific cross-reactivity on Expi293F cell lysate (30 ng/well), BSA (30 ng/well) and lysozyme (30 ng/well). IgGs in the sera were detected using goat-anti-hIgG(Fc)-HRP (1:70,000, A0170, Sigma). 384-well liquid handling was performed with a Precision XS microplate sample processor (BioTek), EL406 washer dispenser (BioTek) and BioStack Microplate stacker (BioTek). OD450 nm-620 nm was measured in an ELISA plate reader (BioTek Instruments, Epoch) and its software Gen5 version 3.03 was used to calculate EC_50_ values, further expressed as relative potency towards an internal calibrant for which the Binding Antibody Unit (BAU) was calculated using the WHO International Standard 20/136 in relation to the original Wuhan strain RBD. The graphics were created by GraphPad Prism 9.1. Significance was calculated by pairwise non-parametric multiple comparison ANOVA (Friedman’s test), using the Wuhan wt RBD values as reference value for all three VOCs, but Omicron data were shown separately for better illustration.

## Data Availability

All data produced in the present study are available upon reasonable request to the authors

## Acknowledgments

We kindly acknowledge the financial support of the European Union for the ATAC (‘‘antibody therapy against corona,’’ Horizon2020 number 101003650), the MWK Niedersachsen (14-76103-184 CORONA-2/20) for the projects ‘‘Antibody generation,’’ ‘‘Neutralization experiments’’ and ‘‘Structure-based analysis of antiviral strategies against CoV-2 target proteins and the Deutsche Herzstiftung (‘‘Menschliche monoklonale Antikörper gegen SARS-CoV2 zur Prophylaxe gegen COVID-19 bei vorerkrankten Risikopatienten – Unterstützung der Entwicklung’’) and the Swedish Research Council. SS was supported by DFG FOR3004. LCS was supported by grants from the Helmholtz Association (EU-Partering PIE-0008 and Helmholtz campaign COVIPA). We would like to thank all the blood donors who agreed to this scientific study.

