## Supplementary Figures for "Human serum from SARS-CoV-2 vaccinated and COVID-19 patients shows reduced binding to the RBD of SARS-CoV-2 Omicron variant"

**Supplementary Figure 1: SDS-PAGE of the RBD25 variants.** 2 µg of the indicated purified RBD25 variant in Laemmli buffer containing 5 % beta-mercaptoethanol were heated to 95°C for 10 min and run on 15% SDS-PAGE.

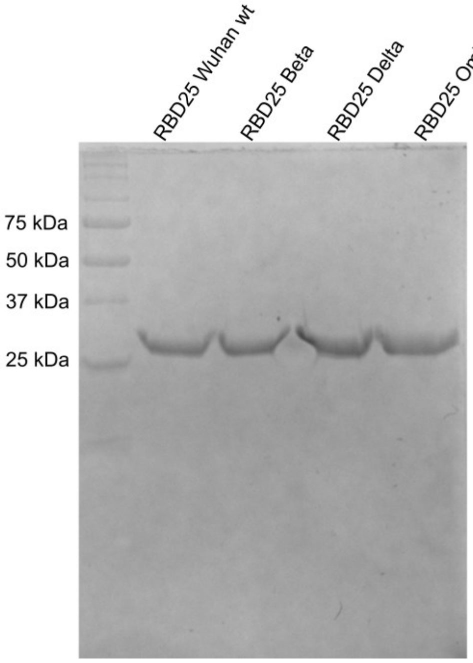


**Supplementary Figure 2: MST measurements of the ACE2-hFc RBD interaction.** Triplicates were measured and Hill fit was applied.

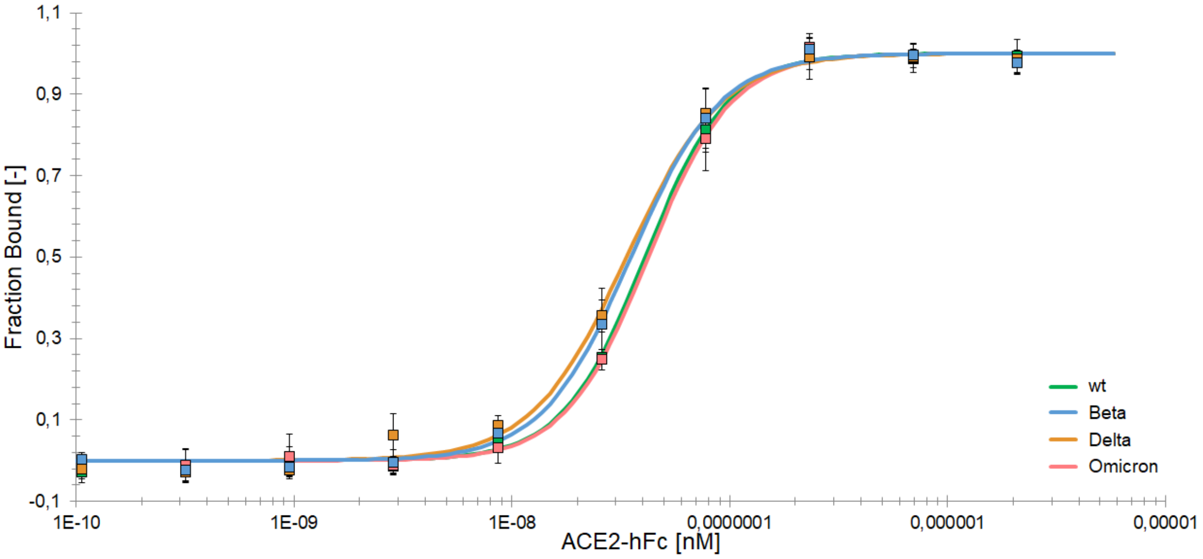
